# Outcomes of COVID-19 Vaccination Efforts in Florida from December 14, 2020 to March 15, 2021 on Older Individuals

**DOI:** 10.1101/2021.04.05.21254722

**Authors:** Scott. A. Rivkees, Shamarial Roberson, Carina Blackmore

## Abstract

Per-capita, Florida ranks second in those 65 years of age and older (20.5%) with more than 4,500,000 individuals in this category. COVID-19 vaccine was allocated in a phased roll-out beginning December 14, 2020. Phase 1A included health care personnel with direct patient contact, and residents and staff of nursing homes (NHs) and assisted living facilities (ALFs). Following this initial phase, individuals 65 years of age and older became eligible for vaccination, along with individuals determined by hospital providers to be extremely medically vulnerable to COVID-19. This strategy was based on the desire to most immediately reduce morbidity and mortality, as COVID-19 morbidity and mortality is age-related. Through March 15, 2021, 4,338,099 individuals received COVID-19 vaccine, including 2,431,540 individuals who completed their vaccination series. Of all those vaccinated, 70% were 65 years of age and older, and 63% of those 65 years of age and older. Beginning February 1, 2021, the decline in the number of new cases per week became greater in those 65 years of age and older than those younger. By March 15, 2021, the number of new cases, hospitalizations, and deaths per day for those 65 years of age and older relative to mid-January, were 82%, 80%, and 92% lower respectively. In comparison, the number of new cases, hospitalizations, and deaths per day for those younger than 65 years of age were 70%, 60%, and 87% lower respectively. Reductions in rates in those 65 year of age and older, were thus greater than in those who were younger (p <0.01; Wilcoxon test). These data show that vaccination efforts directed at those 65 years of age and older results in accelerated rates of overall declines in COVID-19 hospitalizations and mortality.

## Brief Report

Florida is the third most populous state with more than 21,600,000 residents. ^1^ Per-capita, Florida ranks second in those 65 years of age and older (20.5%) with more than 4,500,000 individuals in this category. ^1^ There are more than 3,800 licensed skilled nursing homes (NHs) and assisted living facilities (ALFs) with about 140,000 residents and 150,00 staff.

Through March 15, 2021, 1,954,460 individuals tested positive for COVID-19 in Florida, 82,561 residents have been hospitalized and 32,458 individuals have died from COVID-19. ^2^ Of those hospitalized, 56% were 65 years of age or older. Of those who died from COVID-19, 83% were 65 years of age or older; residents of long-term care facilities (NHs and ALFs) account for 36% of COVID-19 deaths. ^2^

COVID-19 vaccine was allocated in a phased roll-out beginning December 14, 2020. Phase 1A included health care personnel with direct patient contact, and residents and staff of NHs and ALFs. Following this initial phase, individuals 65 years of age and older became eligible for vaccination, per Executive Order 20-315, along with individuals determined by hospital providers to be extremely medically vulnerable to COVID-19. ^3^ This strategy was based on the desire to most immediately reduce morbidity and mortality, as COVID-19 morbidity and mortality is age-related. ^4^

Vaccination of NHs and ALFs involved the Federal Pharmacy Partnership for Long-Term Care Program, and teams from the Florida Division of Emergency Management and the Department of Health. By February 15, 2021, nearly all NHs (99.7%) and ALFs (94%) had residents and/or staff vaccinated. In NHs, 67% of residents and 36% of staff were vaccinated, as were 80% of residents and 36% of staff of ALFs. Vaccination of the 65 years of age and older population began at the end of December 2020, and involved county and state sites, Federally Qualified Health Centers, hospitals, pharmacies, and other community partners. Efforts were also directed at reaching minority and vulnerable populations.

Through March 15, 2021, 4,338,099 individuals received COVID-19 vaccine, including 2,431,540 individuals who completed their vaccination series. ^5^ Of all those vaccinated, 70% were 65 years of age and older, and 63% of those 65 years of age and older were vaccinated, in comparison with 7.5% of those younger than 65 years of age. Of these vaccinated, 6% were Black, 12.6% were Hispanic, and 67% were White.

In Israel, COVID-19 vaccination has been recently observed to result in reduced cases in the general population ^6^, but we are unaware of a similar analysis in the United States. To assess if there was an effect of vaccination efforts focusing on the older population, changes in rates of new cases and hospitalizations were examined relative to the peak number of cases in mid-January for those older and younger than 65 years of age. Beginning February 1, 2021, the decline in the number of new cases per week became greater in those 65 years of age and older than those younger. By March 15, 2021, the number of new cases, hospitalizations, and deaths per day for those 65 years of age and older relative to mid-January, were 82%, 80%, and 92% lower respectively (Figures 1-3). In comparison, the number of new cases, hospitalizations, and deaths per day for those younger than 65 years of age were 70%, 60%, and 87% lower respectively (Figures 1-3). Reductions in rates in those 65 year of age and older, were thus greater than in those who were younger (p <0.01; Wilcoxon test). Collectively, these data show that vaccination efforts directed at those 65 years of age and older resulted in accelerated rates of overall declines in COVID-19 hospitalizations and mortality.

**Figure 1.**
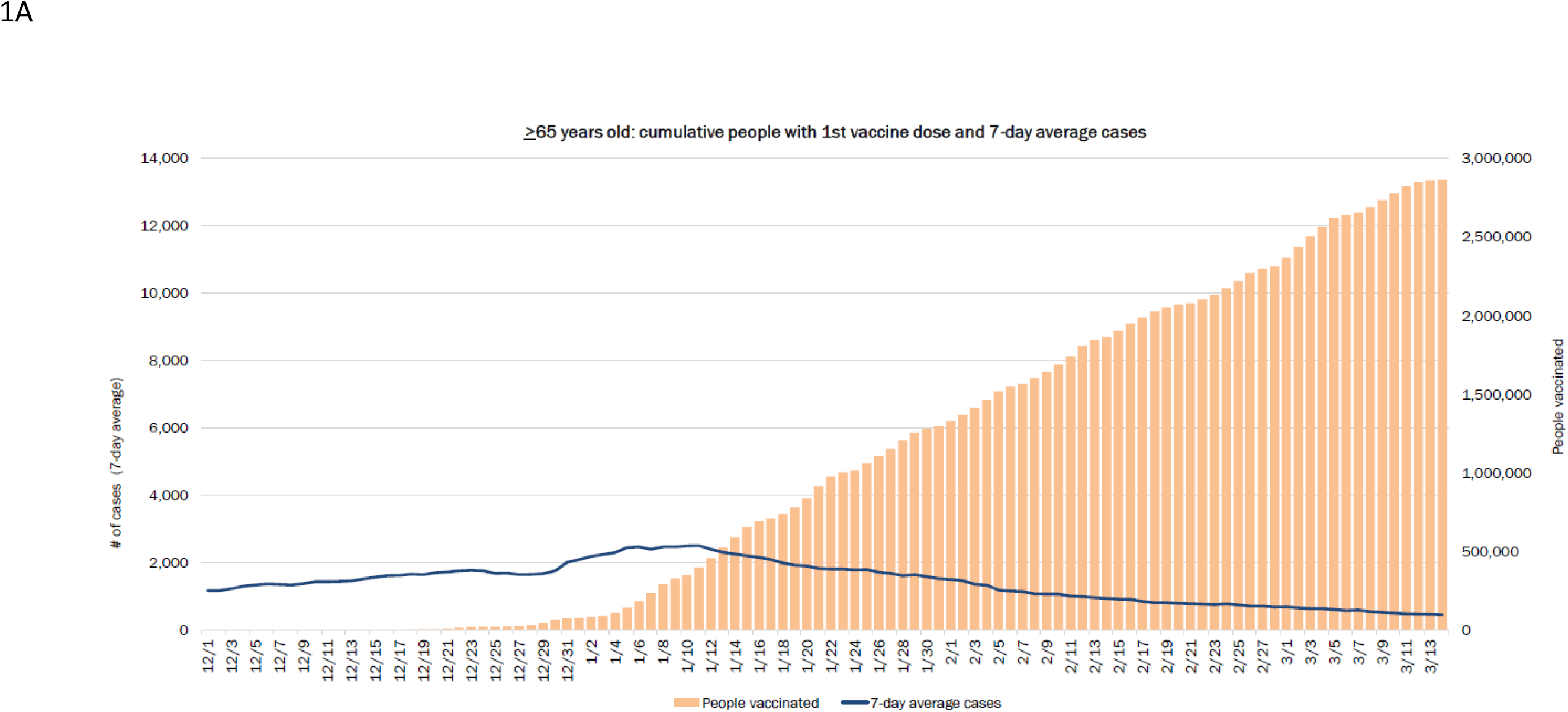
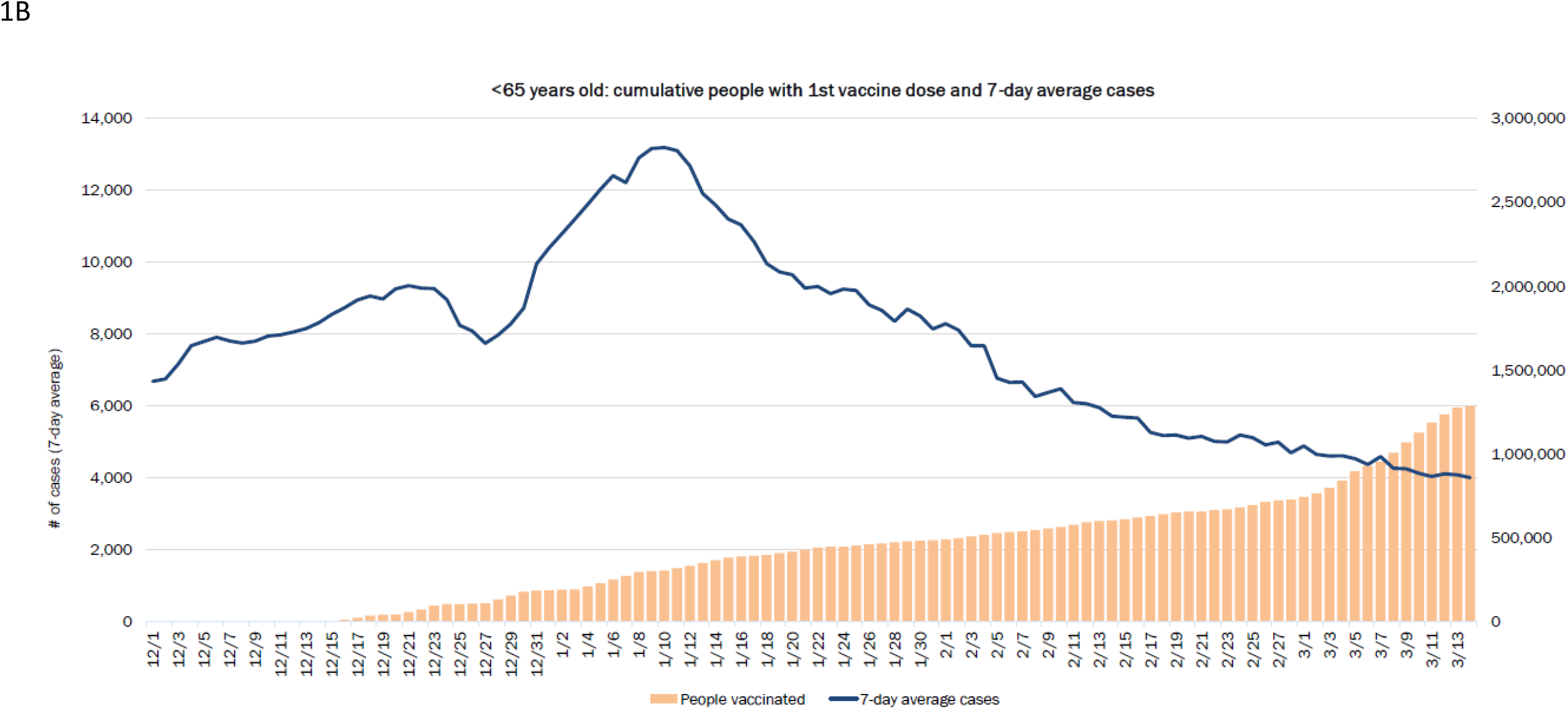
Seven-day rolling averages of cases of individuals (A) 65 years of age and older and those (B) younger than 65 years of age.

**Figure 2.**
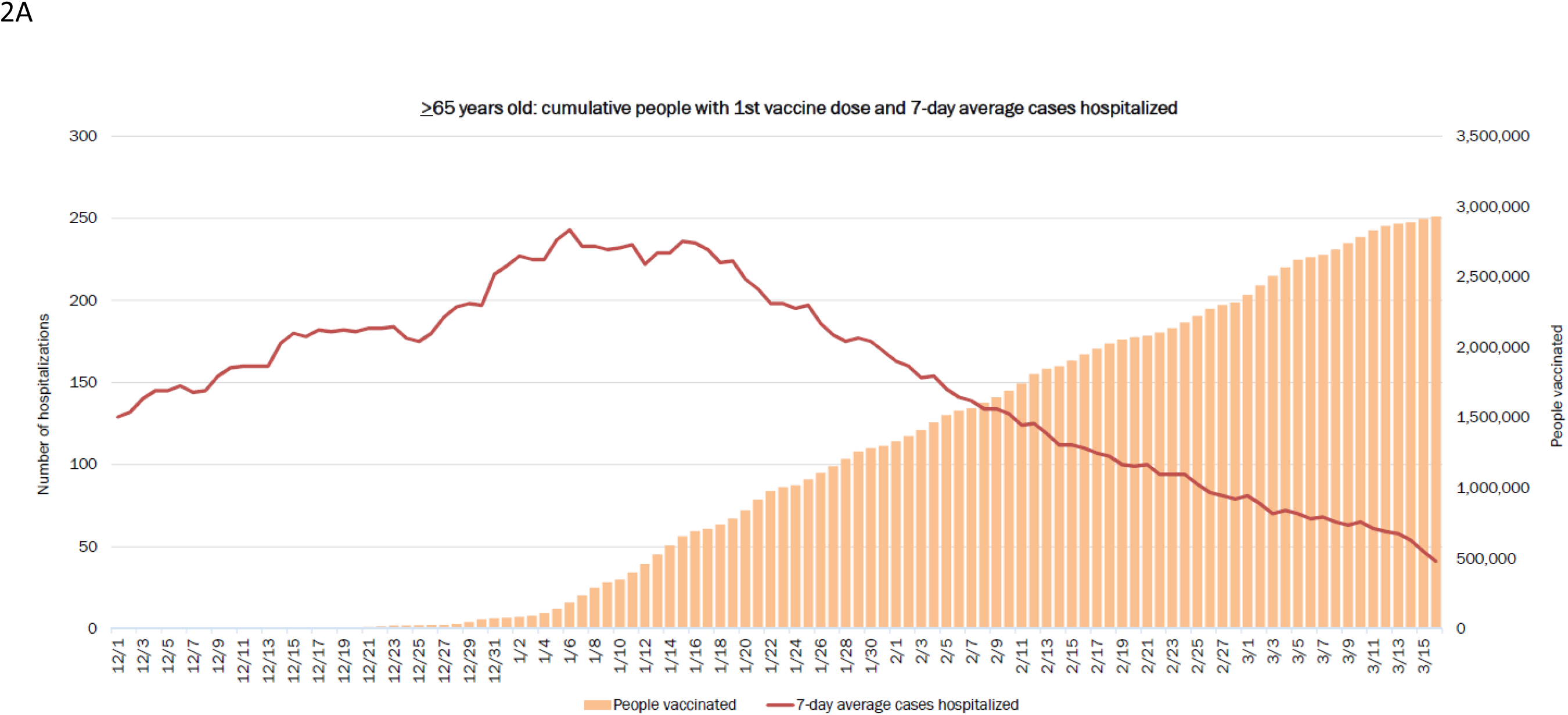
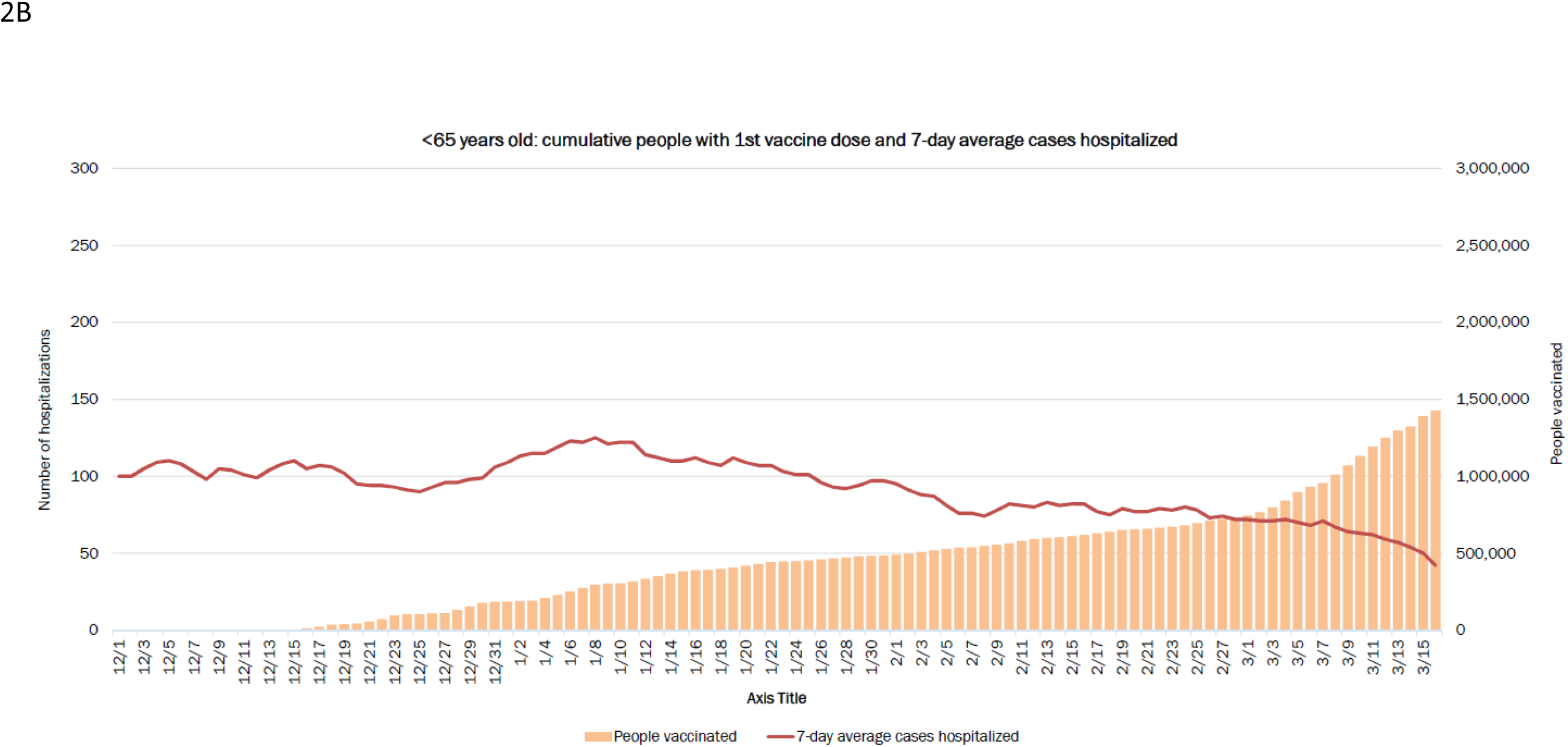
Seven-day rolling averages of hospitalizations of individuals (A) 65 years of age and older and those (B) younger than 65 years of age.

**Figure 3.**
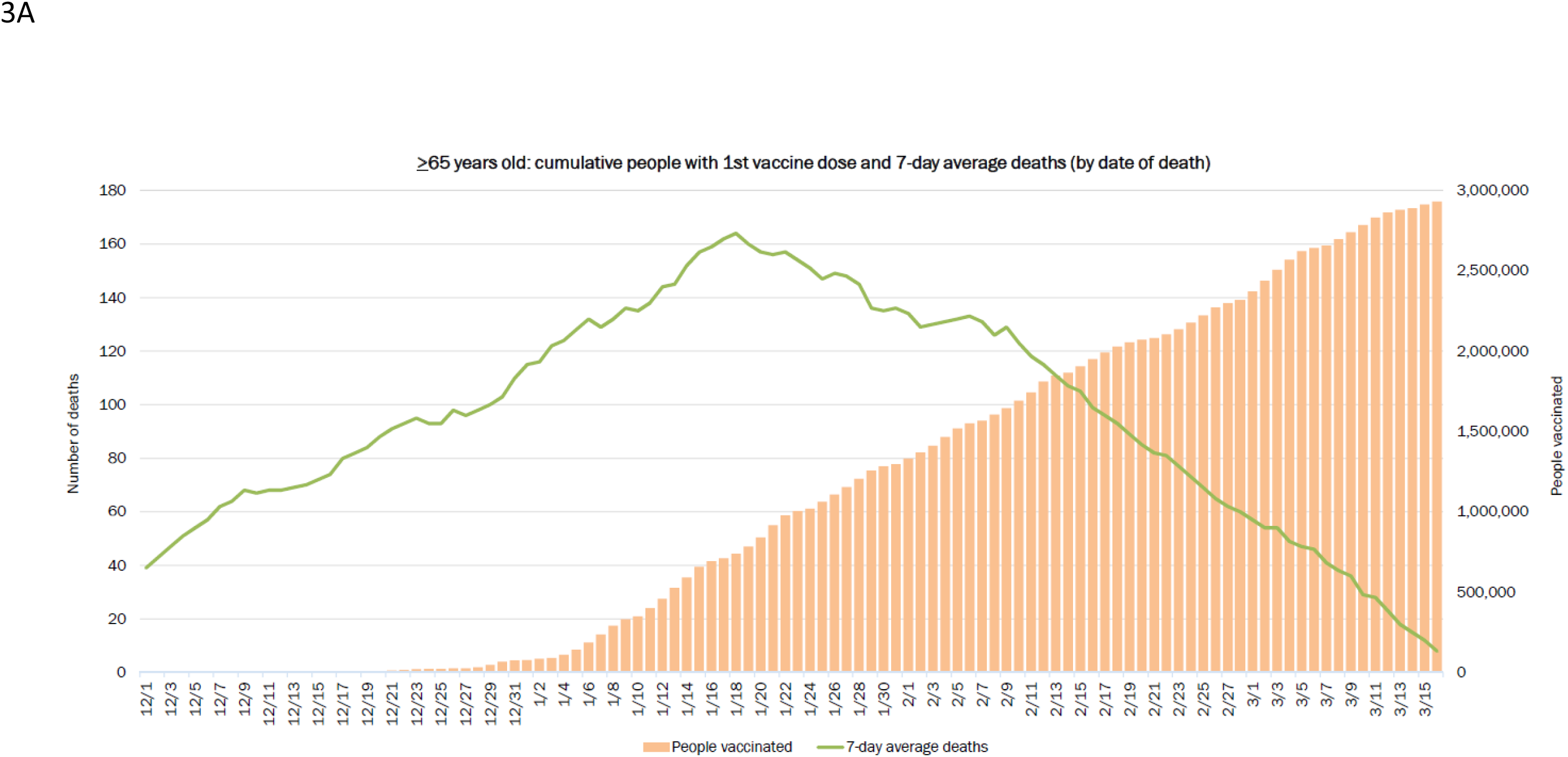
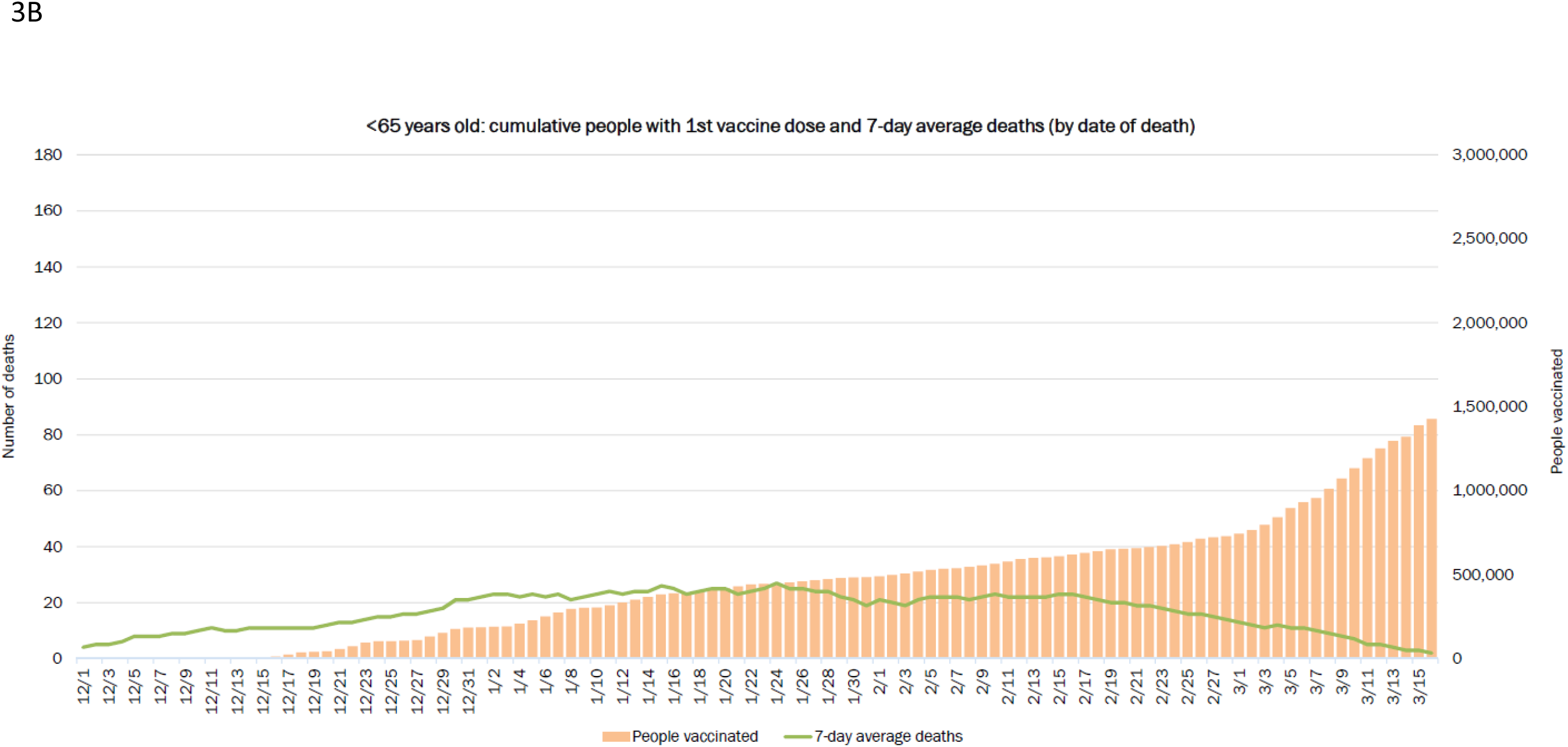
Seven-day rolling averages of deaths of individuals (A) 65 years of age and older and those (B) younger than 65 years of age.

## Data Availability

All data are available in reports posted on the Florida Department of Health website.

http://ww11.doh.state.fl.us/comm/_partners/covid19_report_archive/vaccine/vaccine_report_latest.pdf

